# Clinical and environmental wastewater-based bacteriophage surveillance for high-impact diarrheal diseases, including cholera, in Bangladesh

**DOI:** 10.1101/2025.07.30.25332375

**Authors:** Marjahan Akhtar, Md. Ariful Amin, Subah Nuzhat Hussain, Nazia Nazrul Nafsi, Nasrin Parvin, Farhana Khanam, Md. Taufiqul Islam, Md. Amirul Islam Bhuiyan, Rahima Afroz, Md. Golam Firoj, Fahima Chowdhury, Ashraful Islam Khan, Mohammad Jubair, Edward T. Ryan, B. Jesse Shapiro, Nicholas R. Thomson, Eric J Nelson, Md. Mustafizur Rahman, Yasmin Ara Begum, Taufiqur Rahman Bhuiyan, Firdausi Qadri

## Abstract

Bacteriophages (phages) likely play a critical role in modulating transmission dynamics of diarrheal pathogens. This study investigated the role of phages in modulating the prevalence and seasonal patterns of major diarrheal pathogens, *Vibrio cholerae* O1 (VCO1), enterotoxigenic *Escherichia coli* (ETEC), *Shigella* spp., and *Salmonella* spp. in diarrheal patients and environmental wastewater specimens collected from six different sites in Dhaka, Bangladesh, in 2024. VCO1, ETEC, *Shigella*, and *Salmonella* were detected in 10.1%, 7.8%, 1.7%, and 2.4% of diarrheal specimens, respectively. In contrast, phages targeting these pathogens were more frequently isolated, with detection rates of 20% for VCO1, 30% for ETEC, 57% for *Shigella*, and 9.2% for *Salmonella*-specific phages. Adults showed a significantly higher burden of VCO1 and corresponding phages compared to children <5 years (*P*<0.001). Seasonal analysis revealed significant correlations between VCO1 (37.3%) and corresponding phages (57.6%) peaking in late September in both clinical (R = 0.53, *P*< 0.0001) and environmental wastewater specimens (R = 0.65, *P*<0.001). The highest correlation (R=0.68) was found between the increased rate of wastewater phages in the preceding week and a rise in cholera cases in the following week. ETEC and ETEC phages isolated from wastewater also showed strong correlations (R = 0.65, *P*<0.001). Cross-specificity analysis demonstrated that VCO1 phages were highly specific to their targets, whereas ETEC and *Shigella* phages exhibited broader host ranges, with some *Shigella* phages capable of infecting ETEC and *Salmonella* spp. Overall, these findings underscore the potential of bacteriophages as an alternate or adjunctive tool for cholera surveillance.

**Importance:** Understanding the dynamics between phages and their bacterial hosts is critical for elucidating disease burden, yet their potential for surveillance remains underexplored. To our knowledge, this is the first study that longitudinally investigated major diarrheal pathogens and their phages in both clinical and environmental sources to assess the potential of bacteriophages as a tool to improve diarrheal surveillance. The high frequency of phages compared to the host bacterial counterparts suggests a valuable, yet underutilized, role for phages in surveillance systems. Strong seasonal alignment between *V. cholerae* O1 and its phages, both peaking in late September, suggesting phage dynamics may reflect pathogen transmission. Our findings suggest that wastewater-derived vibrio phages may serve as an early indicator for predicting cholera burden. These results highlight our limited understanding of the complex interplay between phages and their bacterial hosts, particularly in shaping pathogen populations in endemic settings.

## 1. Introduction

Diarrheal diseases remain a leading cause of morbidity and mortality in low- and middle-income countries (LMICs) (1). In Bangladesh, poor sanitation, high population density, and natural disasters contribute to the widespread prevalence of waterborne diseases like diarrhea. The major bacterial pathogens responsible for substantial diarrheal burden in Bangladesh include *V. cholerae* O1, enterotoxigenic *Escherichia coli* (ETEC), *Shigella* spp., and *Salmonella* spp. (2). Continuous surveillance of these high-impact pathogens is important for early outbreak detection, monitoring levels of antimicrobial resistance, guiding vaccine strategies, and identifying seasonal trends that in turn inform public health policies and therapeutics. Traditional disease surveillance tools have primarily focused on detection of these disease-causing pathogens in both clinical and environmental sources (3–5). However, in resource-limited settings, certain environmental challenges like poor water and sanitation infrastructure and the high costs of laboratory diagnostics can hinder effective disease surveillance. Developing a low-cost, accurate, scalable approach could enhance both clinical and environmental surveillance systems. In this context, using bacteriophages as epidemiological markers to track and follow the evolution of specific diarrheal pathogens may offer an alternative or adjunctive option. Virulent bacteriophages (phages) are viruses that infect and subsequently lyse susceptible bacterial hosts. They play an important role in bacterial evolution by serving as natural biocontrol agents as well as vectors facilitating the spread of virulence factors and antibiotic resistance genes through horizontal gene transfer (6, 7). Despite the recognized significance of bacteriophages in bacterial ecology, there is a lack of evidence in the modern era that systematic surveillance programs incorporating phage monitoring alongside traditional pathogen tracking are efficacious.

Understanding environmental and biological factors that influence the seasonal variations of important diarrheal pathogens is critical for improving disease surveillance and developing control strategies. While several environmental factors, such as temperature, rainfall, and water salinity, have been implicated in shaping the seasonal dynamics of diarrheal pathogens (8, 9), the role of bacteriophages in modulating pathogen transmission and evolution remains less studied. In Bangladesh, where cholera or ETEC-associated diarrhea and shigellosis remain endemic, the interactions between bacterial pathogens and their specific bacteriophages could be key to understanding disease seasonality and outbreak dynamics. Studies suggest that environmental bacteriophage populations fluctuate seasonally and may influence the persistence and transmission of diarrheal pathogens in both aquatic environments and human hosts (10, 11). A recent study on *Salmonella* Typhi demonstrated that an increased typhoid burden correlated with a rise in specific bacteriophages in environmental water in that area (12). This finding suggests that bacteriophages could serve as a potential tool for rapid environmental surveillance to assess the risk of typhoid fever and other diseases in the community. In addition, the presence of vibrio phages was shown to hamper cholera diagnostics in the diarrheal patients (13). However, there is limited evidence linking the seasonal prevalence of pathogen-specific bacteriophages with clinical diarrheal cases in endemic settings.

In this study, we aimed to investigate the potential correlations between the seasonal patterns of *V. cholerae*, ETEC, and *Shigella* and *Salmonella* associated diarrhea and the prevalence of pathogen-specific bacteriophages in diarrheal specimens as well as environmental wastewater specimens collected from different sewage sources in Dhaka city, Bangladesh. By analyzing diarrheal cases from a surveillance program in Bangladesh, we sought to determine whether fluctuations in bacteriophage populations correspond to the seasonal variations of these bacterial pathogens.

## 2. Method and materials

### 2.1. Clinical specimens

In Bangladesh, icddr,b Dhaka hospital runs a 2% diarrheal disease surveillance in which every 50^th^ patient admitted to the hospital is enrolled for surveillance purposes. We used this surveillance system to analyze both diarrheal pathogens and corresponding phages from the fecal specimens collected weekly from the enrolled specimens from January to December 2024. A total of 3123 fecal specimens were collected in 2024, including 915 (29%) adults and 2208 (71%) children. Demographic information of the study participants was shown in Table 1. 2% surveillance study is approved by icddr,b Research Review Committee (RRC) and Ethical Review Committee (ERC).

**Table 1:** Demographic characteristics of diarrheal patients.

| Parameters | Diarrheal patients<br>n=3123 |
| --- | --- |
| **Age distribution, n (%)** |  |
| >5 years | 2055 (65.8) |
| 5-17 years | 153 (4.89) |
| >18 years | 915 (29.29) |
| **Sex, n (%)** |  |
| Male | 1761 (56.38) |
| Female | 1362 (43.61) |
| **Dehydration status, n (%)** |  |
| Severe | 443 (14.19) |
| Mild | 652 (20.87) |
| None | 2028 (64.94) |
| **Month wise patient enrollment, n (%)** |  |
| January | 235 (7.52) |
| February | 284 (9.09) |
| March | 298 (9.54) |
| April | 216(6.91) |
| May | 280 (8.97) |
| June | 279 (8.93) |
| July | 226 (7.23) |
| August | 150 (4.80) |
| September | 215 (6.88) |
| October | 257 (8.22) |
| November | 267 (8.55) |
| December | 416 (13.32) |

### 2.2. Environmental wastewater

Environmental wastewater from sewage sources (n=144) was collected from six distinct locations in Dhaka city once every week from July to December 2024. Those sites were selected by the study team by mapping the location and direction of sewage flow. Three of the locations were from the Dakkhinkhan area, and three were from the Mirpur area (Figure 1). Every week, wastewater samples were collected before 9 am in a sterile bottle (Nalgene) and transported to the laboratory. In the laboratory, wastewater samples were settled first and filtered using 0.22 µm Durapore® Membrane filter paper (Merck, Germany). While the filtrates were tested for the presence of bacteriophages, the residues on the filter papers were used for the detection of the bacterial strains.

**Figure 1.**
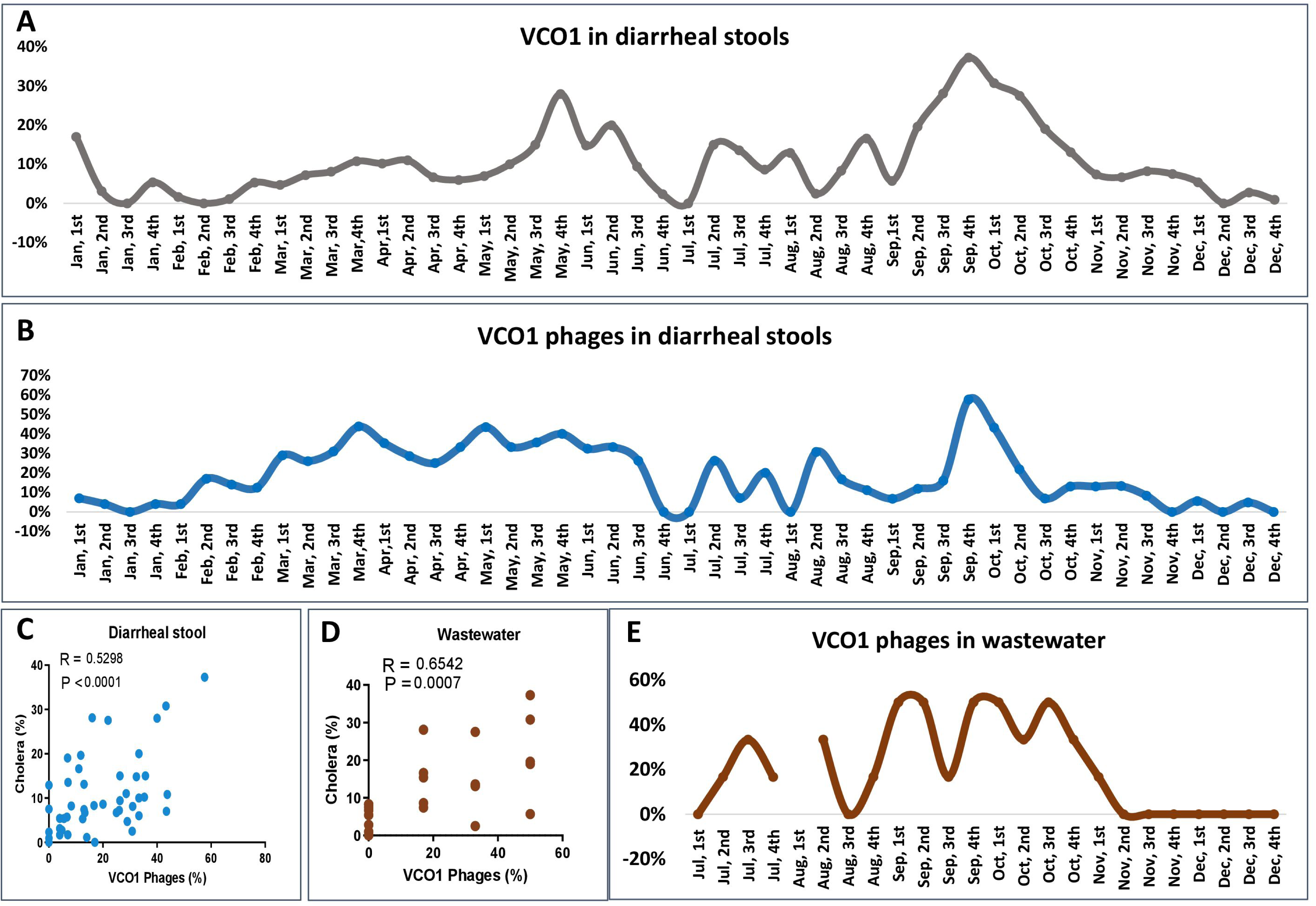
Temporal dynamics of *V. cholerae* O1 (VCO1) and corresponding phages in clinical and environmental specimens. (A) Weekly isolation rate of VCO1 in diarrheal specimens (n=3123) collected between January and December 2024. (B) Weekly isolation rate of VCO1 phages from diarrheal specimens (n=1068) from January to December 2024. (C) Correlation between weekly percentage of cholera cases and phages isolated from diarrheal stools. (D) Correlation between weekly percentage of cholera cases and phages isolated from environmental wastewater specimens (E) Weekly isolation rate of VCO1 phages in environmental wastewater samples collected between July and December 2024. Correlation analyses were performed using the Spearman test.

### 2.3. Identification of diarrheal pathogens

Diarrheal fecal specimens were used to isolate *V. cholerae* O1, ETEC, *Salmonella*, and *Shigella* spp (14–16). For the detection of *V. cholerae*, fecal specimens were streaked onto Tellurite Taurocholate Gelatin Agar (TTGA) and then incubated overnight at 37 °C. An agglutination assay utilizing specific monoclonal antibodies was used to detect *V. cholerae* O1-Ogawa and Inaba serotypes. ETEC was determined using a conventional PCR method targeting heat-labile (LT) and heat-stable (ST) enterotoxins as described previously (3, 15). For the detection of *Shigella* and *Salmonella* spp., biochemical and serological tests were carried out (16).

### 2.4. Host bacterial strains

To screen for bacteriophages, we used 28 different strains as bacterial hosts, including *V. cholerae,* ETEC, *Shigella*, and *Salmonella* spp. (supplementary Table 1). For *V. cholerae,* phages were isolated using two different *V. cholerae* O1 serotypes: Ogawa and Inaba. In addition, we used a recombinant *V. cholerae* O1 strain (AC-6169, *V. cholerae* E7946 ΔRS1-CTXphage-TLC, ΔK139-attB, wbeL-A111G, A114G, manAA216G, A219G, A609G, A612G; a generous gift from Dr. Andrew Camilli at Tufts University care of Drs. M. Alam (icddr,b) and E. Nelson (University of Florida). To identify ETEC-specific phages, we utilized 12 different ETEC strains without or with known colonization factors (CFA/I, CS1+CS3+CS21, CS2+CS3+CS21, CS5+CS6, CS4+CS6, CS6+CS8, CS7+, CS12+, CS14+, CS17+, LT+CF-, and ST+CF-ETEC strains). *Shigella*-specific phages were screened against four different *Shigella* spp.: *Shigella flexneri*, *Shigella sonnei*, *Shigella boydii*, and *Shigella dysenteriae*. Similarly, *Salmonella* phages were identified against *Salmonella enterica* serovars Typhi, Paratyphi A, and non-typhoidal *Salmonella* strains, including *S. enterica* group B and group C1. Most of the host bacterial strains (n=19) were characterized using whole genome sequencing as described in the below section. An antibiogram assay was performed for determination of antibiotic resistance as described previously (17) and results are shown in Supplementary Table 1.

### 2.5. Whole genome sequencing (WGS) and bioinformatics analysis

WGS was performed in the host bacterial strains. After overnight culture in lauria broth, DNA extraction was carried out using the Qiagen DNeasy Blood & Tissue Kit. The extracted DNA’s quality was evaluated for suitability for subsequent whole genome sequencing (WGS) using both Nanodrop (Thermo Fisher Scientific) and Qubit (Thermo Fisher Scientific) measurements. A Nanodrop spectrophotometer was used to assess DNA purity, where a 260/280 reading of 1.8 indicated high quality and minimal contamination. DNA quantification was performed using a Qubit 4.0 Fluorometer, yielding a good amount of concentration. Library preparation was performed using Illumina DNA Prep Reagent Kit and epMotion 5075. Prepared DNA libraries were sequenced using the Illumina NextSeq2000 platform. The quality of these raw sequencing reads was assessed using FastQC (v0.12.1). Trimming was done to remove any adapters and low-quality sequences, employing tools such as BBDuk (v39.01) and Fastp (v0.23.4). The high-quality, trimmed reads were then ready for the subsequent analysis steps. High-quality, trimmed paired-end reads were assembled de novo into contigs using the SPAdes genome assembler (v4.1.0). The reads were screened for the presence of antimicrobial resistance (AMR) genes using Ariba (v2.14.6). Ariba was run against the Comprehensive Antibiotic Resistance Database (CARD) to identify known AMR genes and their associated resistance mechanisms. AMR data are shown in supplementary Table 2. To identify putative bacteriophage defense systems within the assembled genomes, the contigs were analyzed using DefenceFinder (v2.0.0). Phage resistance genes are shown in supplementary Table 3. To predict the presence of virulence genes linked to ETEC, specifically to toxins (LTh, STh, and STp) and colonization factors, the ETEC virulence and CF database employed in this study were sourced from Astrid von Mentzer’s repository, accessible at https://github.com/avonm/.

### 2.6. Identification of phages specific to diarrheal pathogens

Bacteriophages against ETEC, *V. cholerae, Shigella*, and *Salmonella* spp. were screened using a plaque assay (18, 19). Host bacterial strains were grown in Luria broth at 37°C with shaking (200 rpm) until mid-log phase (OD600 = 0.4-0.6). Bacterial lawns were prepared by spreading 1 ml of culture onto Luria Agar plates and drying under aseptic conditions. This drying step ensured that phage samples were not overlapped upon spotting, allowing for clear formation of plaques. Fecal samples were diluted ten-fold prior to centrifugation (10,000 x g for 10 minutes). Meanwhile, water samples were filtered (0.22 µm) to remove debris and enriched with respective bacterial hosts (overnight in LB at 37°C) before serial dilution. Finally, supernatants were spotted onto bacterial lawns and incubated at 37°C overnight. Clear plaques indicated phage-mediated bacterial lysis, confirming the presence of lytic bacteriophages.

### 2.7. Phage purification and cross-specificity determination

Phages identified in the plaque assay were subsequently isolated and purified to obtain single-phage populations. Single plaques were picked, diluted in PBS, and re-spotted on fresh bacterial lawns. This process was repeated at least four times to ensure the purity of the isolated phages. Purified phage lysates were stored at 4°C for short-term use and at -80°C with 50% glycerol for long-term preservation.

### 2.8. Statistical analysis

Descriptive statistics for all the demographic and clinical characteristics are presented as frequencies and percentages. The Spearman correlation test was used to evaluate seasonal patterns of weekly isolated bacteria and phages. The chi-square test was used to measure the difference between the burden of diarrheal pathogens and corresponding phages in different age groups of patients. All statistical tests have been conducted at the 5% level of significance. Data was analyzed by using Excel, GraphPad Prism (version 6.0), and R (version 4.4.2). Time-lagged cross-correlation analysis was conducted using the “forecast” package from R.

## 3. Results

### 3.1. Diarrheal pathogens and phages

Among the 3,123 diarrheal specimens (fecal and rectal swab) tested, *V. cholerae* O1 was detected in 314 (10.1%), ETEC in 244 (7.8%), and *Shigella* spp. in 52 (1.7%) of stool specimens, while *Salmonella* spp. was identified in 2.4% of specimens (Table 2). The overall burden of bacteriophages was significantly higher than their corresponding diarrheal pathogens in the fecal specimens (Table 2, *P* <0.0001). Among the diarrheal specimens tested for phages (n=1,068), 20% were positive for *V. cholerae* O1-specific phages, 30.8% for ETEC phages, 47.1 % for *Shigella* phages, and 5.2% for *Salmonella* phages. Phages isolated on *V. cholerae* O1 hosts could infect both Ogawa and Inaba serotypes. The most abundant ETEC phages were those targeting CS4+CS6, CS1+CS3+CS21, CS2+CS3+CS21, CS12, CS7, CS17, and CS14. Although ETEC strains carrying CFA/I and CS5+CS6 are the most prevalent types circulating in Bangladesh, phages specific to these strains were less frequently detected. Among the isolated *Shigella* phages, those targeting *Shigella sonnei* were the most abundant (40%), followed by *Shigella flexneri* (27%), *Shigella boydii* (13%), and *Shigella dysenteriae* (13%) specific phages. *Salmonella* phages (9.2%) in diarrheal specimens were primarily detected against *Salmonella* Paratyphi A (34 out of 56 *Salmonella* phages), followed by non-typhoidal *Salmonella*, including *Salmonella* serogroups B and C1 and *Salmonella* Typhi (Table 2).

**Table 2:** Isolation rate of *V. cholerae* O1, ETEC and *Shigella* bacteria and corresponding phages isolated from diarrheal patients.

|  | Bacterial pathogens,<br>n (% , CI) | Phages,<br>n (% , CI) | P value |
| --- | --- | --- | --- |
| **Number of tested specimens** | 3123 | 1068 |  |
| *V. cholerae* O1 | 314 (10.1, 9.1-11.1) | 215 (20.1, 17.8-22.7) | <0.0001 |
| ETEC | 244 (7.8, 6.9-8.8) | 329 (30.8, 28.0-33.7) | <0.0001 |
| *Shigella* spp. | 52 (1.6, 1.2-2.2) | 503 (47.1, 44.1-50.1) | <0.0001 |
| *Salmonella* spp. | 76 (2.4, 1.9-3.0) | 55 (5.15, 3.9-6.6) | <0.0001 |

### 3.2. Temporal dynamics of *V. cholerae* O1 and phages in clinical and environmental sources

We analyzed the prevalence of *V. cholerae* O1 and its associated phages in diarrheal stools and wastewater sources over time to investigate their seasonal dynamics. In diarrheal specimens, the proportion of *V. cholerae* O1-positive samples varied throughout the year (Figure 1A). The prevalence remained low during the earlier months but exhibited a gradual increase starting in May and declined in first week of July and remained low until mid-August. In late September, cholera detection rate increased to its highest level (37.3%) before declining towards the end of the year. Similarly, phages targeting *V. cholerae* O1 in diarrheal stools followed similar fluctuating patterns of *V. cholerae* O1 prevalence (Figure 1B). Phage detection frequencies gradually increased from the beginning of the year, with peaks corresponding to late summer (May-June) and early autumn months. Notably, a sharp increase in the number of samples with detectable *V. cholerae* O1 phages (57.6%) was observed around late September, coinciding with the peak *V. cholerae* O1 host detection in fecal specimens. Environmental water samples, analyzed between July and December, also revealed periodic fluctuations in *V. cholerae* O1 phages (Figure 1E). The phage prevalence followed a seasonal trend, peaking multiple times between July and October before reducing towards the end of the year. Using culture-based approaches to detect *V. cholerae* O1 in environmental water specimens had been challenging in this study. However, from mid-August to September, when cholera rate was highest, 30-40% water samples were found *V. cholerae* O1 positive in that period (Supplementary Figure 1).

We further analyzed the correlation between the weekly detection of *V. cholerae* O1 phages, isolated from both diarrheal specimens and wastewater samples, and *V. cholerae* O1 positive diarrheal cases. A statistically significant positive correlation was observed between the presence of *V. cholerae* O1 bacteria and *V. cholerae* phage in diarrheal stools (R = 0.53, *P*<0.0001; Figure 1C). Similarly, *V. cholerae* O1 phage detection in water samples also correlated significantly with the rate of cholera (R = 0.65, *P* = 0.0007; Figure 1D).

We also performed a time-lagged correlation analysis to investigate the temporal association between the presence of *V. cholerae* O1-specific phages in stool or wastewater specimens and the proportion of cholera cases in preceding and subsequent weeks (Figure 2). For stool-derived phages, the strongest correlation (R=0.51) was found for co-occurrence of the presence of phages and proportion of cholera cases during the same week (Figure 2A). Interestingly, for wastewater-derived phages, the highest correlation (R=0.68) was found between phage detection in wastewater samples one week prior and an increased proportion of cholera cases in the following week (Figure 2B). These findings suggest there is a potential application of vibriophages in cholera disease surveillance.

**Figure 2.**
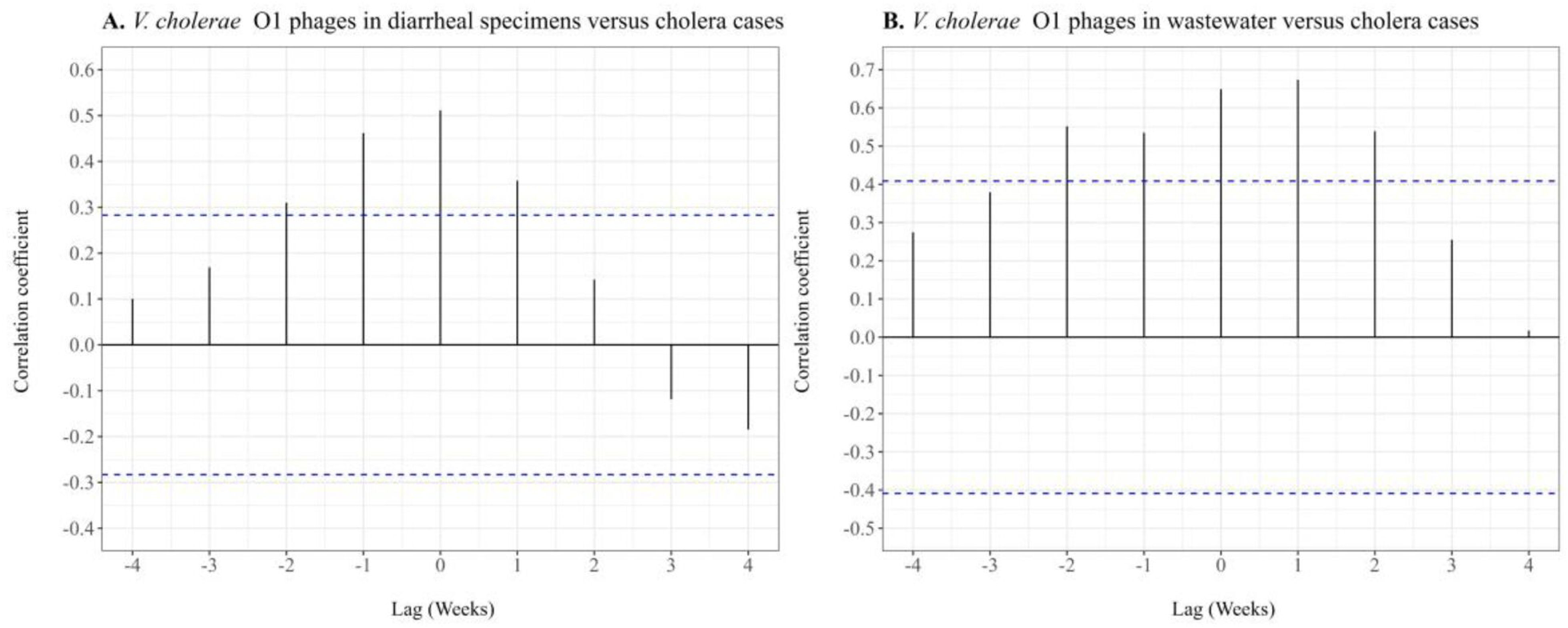
Time-lagged correlation analyses of *V. cholerae* O1-specific phages and cholera cases. (A) Correlation between the percentage of weekly isolation of *V. cholerae* O1 phages in diarrheal stool specimens and cholera cases in preceding and subsequent weeks. (B) Correlation between the percentage of weekly detection of *V. cholerae* O1 phages in environmental wastewater samples and cholera cases in preceding and subsequent weeks. Positive lag weeks represent phage detection preceding cholera cases, while negative lags represent phage detection following cholera cases. Spearman correlation coefficients were calculated for each lag week. The blue horizontal line indicates the 95% significance threshold (p < 0.05), suggesting statistically significant correlations above this line.

### 3.3. Seasonal variation of ETEC and ETEC phages in clinical and environmental sources

The temporal analysis of ETEC and corresponding phages in diarrheal stools showed the detection rate for ETEC in diarrheal stool fluctuated between 0% and 17%, whereas ETEC phage prevalence was higher, ranging between 4% and 75% (Figure 3A). The ETEC phage showed periodic peaks, with the highest prevalence observed between March and May, followed by another increase towards the end of the year. Correlation analysis between ETEC and ETEC phage presence in fecal samples indicated no association (R = 0.08, Figure 3B), suggesting that ETEC phage abundance in diarrheal samples may not directly reflect ETEC burden in infected individuals.

**Figure 3.**
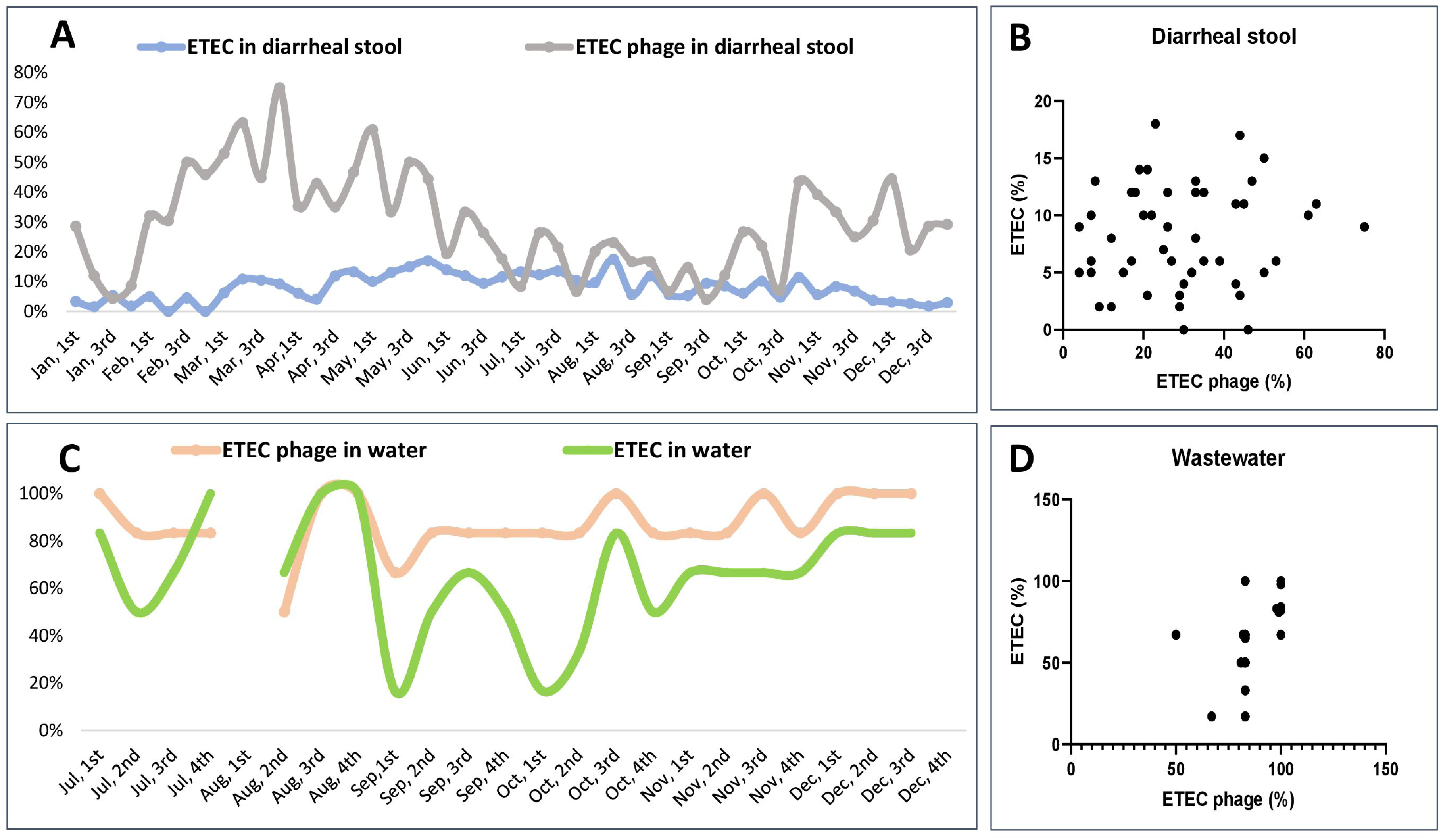
Temporal dynamics of ETEC and corresponding phages in clinical and environmental specimens. (A) Weekly isolation rate of ETEC and ETEC phages from diarrheal specimens collected between January and December 2024. (B) Correlation between the weekly percentage of ETEC and its phages isolated from diarrheal stools. (C) Weekly isolation rate of ETEC and ETEC phages from environmental wastewater specimens collected between July to December 2024. (C) Correlation between weekly percentage of ETEC and its phages isolated from environmental wastewater specimens. Correlation analyses were performed using the Spearman test.

Similarly, ETEC and ETEC phages demonstrated substantial seasonal variation from environmental water samples between July and December, (Figure 3C). The prevalence of ETEC phages in water is consistently high, often exceeding 80%, while ETEC detection fluctuates significantly, with distinct declines in September and October. ETEC isolated from diarrheal stool did not follow a similar pattern, as such a high burden was found in ETEC from wastewater sources (R=-0.01, data not shown). However, there is a strong positive correlation between ETEC and ETEC phage abundance in water sources (Figure 3D; R = 0.65, *P*<0.001), indicating a significant relationship between bacterial and phage presence in the environment.

### 3.4. Age-specific burden of diarrheal pathogens and phages

Enteric infections with *V. cholerae*, ETEC, *Shigella*, and *Salmonella* exhibit age-specific distributions; for example, *V. cholerae* infections are more common in adults than children (5). We were interested in analyzing whether phages specific to these pathogens also follow similar age-related patterns. The distribution of *V. cholerae* O1 bacteria and phages among diarrheal cases was analyzed across three age groups: 0–5 years, >5–17 years, and ≥18 years (Figure 4). Within all diarrheal patients studied, the presence of *V. cholerae* O1 was significantly higher among adults (≥18 years), with approximately 8% of diarrheal cases testing positive, compared to 2% in children aged 0–5 years and a slightly lower percentage in the 5–17 years group (*P* < 0.0001, Figure 4A). A similar trend was observed for *V. cholerae* O1 phages, where their prevalence was markedly higher in the adult group (≥18 years), reaching nearly 18% of diarrheal cases, compared to around 4% in both the 0–5 years and >5–17 years groups (*P* < 0.0001, Figure 4B). These findings suggest a significant age-dependent association between *V. cholerae* O1 and its associated phages, with the highest detection of both bacterial host and phages seen in adults. However, phages targeting ETEC, *Shigella*, and *Salmonella* spp. did not follow any age-specific pattern (data not shown).

**Figure 4.**
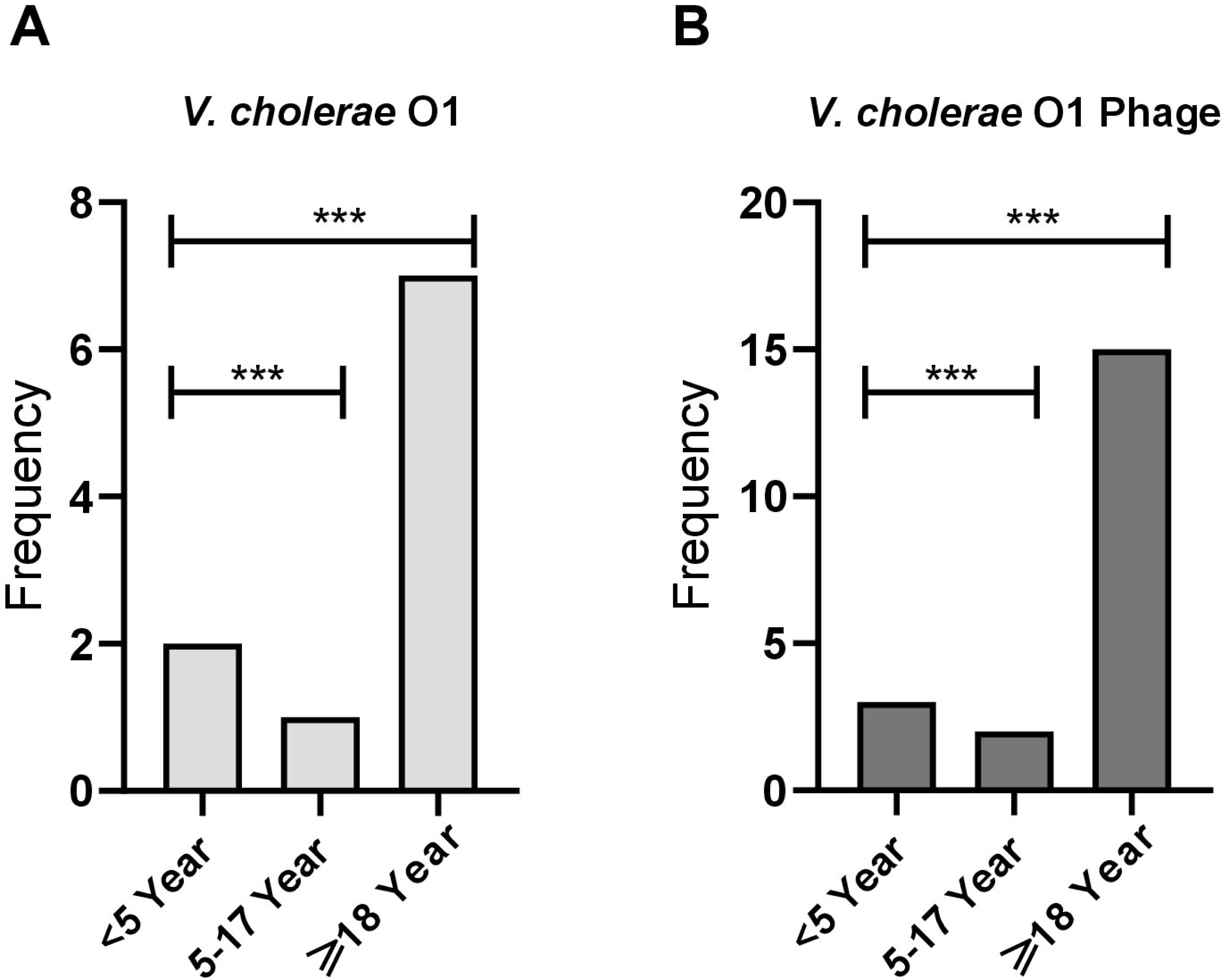
Age-specific burden of *Vibrio cholerae* O1 bacteria (A) and phages (B) in diarrheal specimens. For statistical analysis, chi square tests were performed between <5 years age group versus 5-17 years or ≥18 years. ***P<0.0001.

### 3.5. Cross-specificity of phages

A subset of the purified phages isolated on one pathogen were tested for broad host ranges to 30 different bacterial host strains. *V. cholerae* O1 phages exhibited strict specificity for *V. cholerae* O1 but produced plaques on both the Ogawa and Inaba serotypes. However, *V. cholerae* O1 phages did not display cross-specificity toward *V. cholerae* O139, non-O1/O139 vibrios, or other bacterial species, including ETEC, *Salmonella* spp., and *Shigella* spp. (Figure 5).

**Figure 5.**
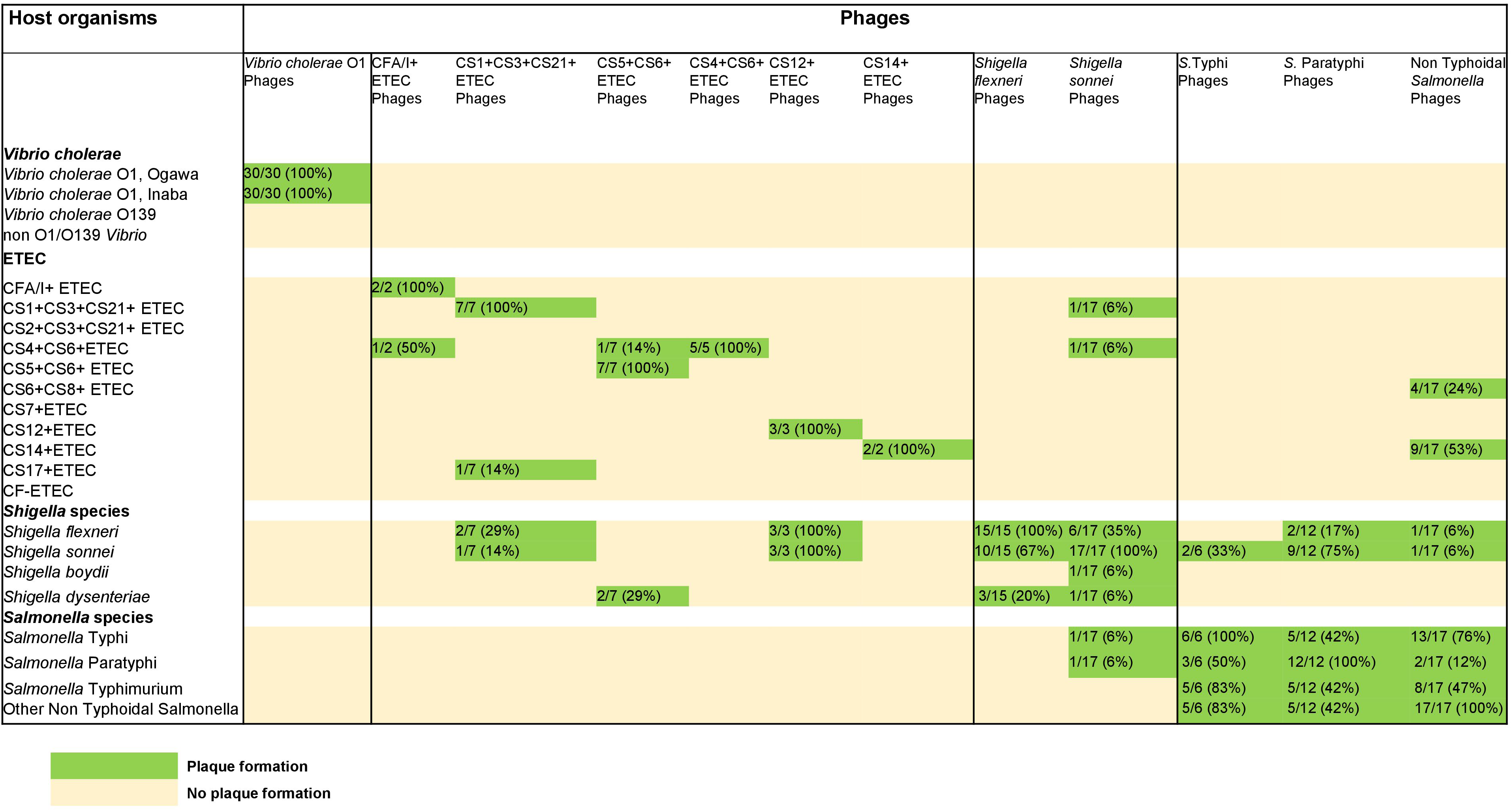
Host ranges of phages isolated against *Vibrio cholerae*, ETEC, *Shigella* spp., and *Salmonella* spp.

ETEC-specific phages demonstrated selective specificity for distinct colonization factor (CF)-positive ETEC strains. For example, phages targeting CS4+CS6+, CS14+ and CS12+ ETEC strains failed to form plaques on other CF-positive ETEC strains. However, phages specific to CFA/I+, CS1+CS3+CS21+ and CS5+CS6+ ETEC strains exhibited limited cross-infectivity with a subset of other CF-positive ETEC strains. Additionally, CS1+CS3+CS21+, CS5+CS6+, and CS12+ ETEC phages displayed broad host ranges towards *Shigella flexneri*, *Shigella sonnei*, and *Shigella dysenteriae* but did not plaque on *V. cholerae* or *Salmonella* spp. (Figure 5).

*Shigella flexneri* phages were capable of infecting all major serotypes, including *S. flexneri* 2a, 3a, and 6. However, these phages also exhibited cross-specificity toward *Shigella sonnei* and *Shigella dysenteriae*. A few phages isolated on *Shigella sonnei* showed a broad host ranges plaquing on *S. flexneri*, *Shigella boydii and Shigella dysenteriae* isolates as well as ETEC. One of the *Shigella sonnei* phages also plaqued on *Salmonella* Typhi and *Salmonella* Paratyphi hosts. None of the *Shigella* phages infected any tested strains of *V. cholerae*. Salmonella phages showed broad host specificity towards different species of *Salmonella*, *Shigella*, and ETEC (Figure 5).

## Discussion

Our study provides insights into the prevalence and dynamics of diarrheal pathogens and their corresponding bacteriophages in fecal and environmental wastewater specimens collected from Dhaka city, Bangladesh. To our knowledge, this is the first study to simultaneously investigate major diarrheal pathogens and their phages in both clinical and environmental sources longitudinally over time to assess the potential of bacteriophages as a tool to improve diarrheal disease surveillance. *V. cholerae* O1 phages are of particular interest and might emerge as a promising tool for cholera surveillance in both clinical and environmental settings. In Dhaka, Bangladesh, *V. cholerae* O1 and ETEC typically exhibit distinct biannual seasonal patterns, with two peaks occurring before and after the monsoon season. The first surge begins in the spring, at the onset of the hot season, while the second peak emerges in autumn, following the monsoons (20–23). Temporal analysis of *V. cholerae* O1 and its phages demonstrated significant seasonal alignment, with peak cholera prevalence occurring in late September, followed by an increase in *V. cholerae* O1-specific phages. A strong correlation between *V. cholerae* O1 phages and bacterial presence in both diarrheal stools and environmental water suggests a dynamic equilibrium, where phage abundance fluctuates in response to bacterial prevalence (23, 24). We also observed a strong correlation between the increased rate of wastewater phages in the preceding week and a rise in cholera cases in the following week and vice versa. Our findings suggest that wastewater-derived vibrio phages may serve as an early indicator for predicting cholera burden. A previous study has shown that environmental phages may influence the occurrence of epidemics and cholera seasonality (24). However, that study reported an inverse correlation between the environmental concentration of vibrio phages and the presence of susceptible *V. cholerae* strains in the water samples collected from lakes or rivers. In contrast, our findings demonstrate a strong positive correlation, where an increase in cholera burden was associated with a rise in *V. cholerae* O1 phages in both clinical and wastewater samples collected from different sewage sources in Dhaka city.

In resource-poor settings, wastewater sampling, which commonly contain fecal matter, is an essential tool for public health surveillance, allowing the tracking of diseases caused by fecal-oral transmitted pathogens, as well as respiratory viruses such as SARS-CoV-2 (25, 26). Notably, we were unable to isolate culture-positive *V. cholerae* O1 from water samples throughout the year, except during the peak epidemic period. Due to limited resources, we were unable to test water samples by molecular techniques such as PCR, which could have detected *V. cholerae* O1 more sensitively (27). PCR-based detection requires well-equipped laboratory and skilled personnel, making it less feasible for resource-limited field settings. Alternatively, given the minimal resources required for environmental phage surveillance, cholera phage detection using a simple plaque assay offers a practical and cost-effective approach. This method can be readily implemented in field settings across endemic regions like Bangladesh and can serve as a valuable complement to existing cholera surveillance strategies as well as to evaluate cholera vaccine effectiveness.

In addition to seasonal trends, we also observed age-specific correlations in cholera burden and phage abundance in stool. Age-stratified analysis revealed a significantly higher prevalence of *V. cholerae* O1 and its associated phages among adults (≥18 years) compared to younger age groups. This finding mirrors the established age-specific cholera burden observed in previous cholera surveillance studies in Bangladesh, which reported higher prevalence among adults than in younger populations (5, 28, 29). A similar trend was observed for *V. cholerae* O1 phages, suggesting that adults may be more frequently exposed to *V. cholerae* O1. Furthermore, the severity of diarrheal disease in cholera patients has been shown to be characterized by the phage-to-bacteria ratio in the human gut (30). All of these findings underscore the intricate interactions between bacterial pathogens and phages, highlighting their potential role in shaping disease epidemiology and transmission dynamics.

Phages targeting ETEC, *Shigella*, and *Salmonella* species were more frequently detected than their corresponding bacterial hosts in diarrheal stools but did not follow seasonal patterns of variation like cholera phages in stools. By contrast, a significant positive correlation was observed between ETEC and its phages in water sources, implying that phage abundance may be influenced by bacterial availability in the aquatic environment. One possible explanation for the high abundance of these bacteriophages in stool and environmental water is their potential role in shaping bacterial populations, either by reducing pathogen loads in the human gut or by persisting in environmental reservoirs until favorable conditions arise for bacterial proliferation (Weinbauer, 2004; Seed, 2015). Our findings also suggest ETEC phages could be specific to colonization factors (CFs); phages specific to certain CF families (CFA/I+, CS1+CS3+CS21+, and CS6+CS8+ETEC) exhibited limited cross-infectivity with a subset of other CF-positive ETEC strains. These findings highlight the need for further investigation of these ETEC phages to find more prevalent ETEC CF-specific phages from the clinical and environmental settings.

Consistent with our findings, previous studies have also reported a high prevalence of *Shigella*-specific phages in various environmental samples, including wastewater (31, 32). Another possible factor contributing to the high abundance of ETEC- and *Shigella*-specific phages is the observed cross-specificity of certain phages, which enables them to infect both bacterial species. This adaptability may facilitate their persistence in the human gut by utilizing alternative hosts. Given that ETEC and *Shigella* share a common evolutionary lineage within the family *Enterobacteriaceae*, their phages may exhibit broad host ranges, a phenomenon that has been documented in different studies (31, 33). Our findings suggest that phages with a narrow host range show stronger correlations with the presence of their target pathogens, while those with a broader host range exhibit weak or no correlations. Despite the presence of resistance genes identified through genomic analysis of host bacterial strains, phages were able to infect and lyse these bacteria. This suggests that phages may possess adaptive mechanisms to overcome the bacterial defense systems. However, due to limited resources, we were unable to perform genomic sequencing of the phages to elucidate the underlying mechanisms of this evasion. Future studies should focus on elucidating the mechanisms underlying phage-host interactions, particularly the factors governing phage specificity and cross-infectivity. This study highlights the intricate relationship between diarrheal pathogens and their corresponding phages, demonstrating significant seasonal variations, age-related patterns, and potential cross-infectivity. Longitudinal studies incorporating metagenomic analyses could provide deeper insights into the role of phages in bacterial evolution as well as antibiotic resistance dissemination. Overall, the findings of this study underscore the potential to integrate phage surveillance into diarrheal disease monitoring programs and provide a foundation for future research into phage-based interventions for enteric infections.

## Data availability

The whole genome sequence data of the host bacteria used to isolate phages have been submitted to the Sequence Read Archive (SRA) under the BioProject number PRJNA1295879. All the metadata linked to these strain numbers of each read-pair are available at http://www.ncbi.nlm.nih.gov/bioproject/1295879. Accession numbers of each read-pair sequence data are shown in Supplementary Table 4.

## Author Contributions

FQ, MA and TRB designed and planned the studies. MA, YAB, MAA, SNH, NNN, NP and MGF performed laboratory experiments and data analyses. MA wrote the manuscript. FQ, TRB, FK, MTI, MAIB, RA, MJ, FC, AIK, ETR, NRT, JS, EJN and MMR reviewed and revised the manuscript. All authors contributed to the interpretation of results and critical review and revision of the manuscript and have approved the final version.

## Acknowledgments

We acknowledge the support of the study participants as well as the dedicated hospital, field, and laboratory workers in this study at icddr,b.

## References

1. Estimates of the global, regional, and national morbidity, mortality, and aetiologies of diarrhoea in 195 countries: a systematic analysis for the Global Burden of Disease Study 2016. The Lancet Infectious diseases. 2018;18(11):1211–28.

2. Kotloff KL, Nataro JP, Blackwelder WC, Nasrin D, Farag TH, Panchalingam S, et al. Burden and aetiology of diarrhoeal disease in infants and young children in developing countries (the Global Enteric Multicenter Study, GEMS): a prospective, case-control study. Lancet (London, England). 2013;382(9888):209–22.

3. Akhtar M, Begum YA, Isfat Ara Rahman S, Afrad MH, Parvin N, Akter A, et al. Age-dependent pathogenic profiles of enterotoxigenic Escherichia coli diarrhea in Bangladesh. Front Public Health. 2024;12:1484162.

4. Khanam F, Islam MT, Bhuiyan TR, Hossen MI, Rajib MNH, Haque S, et al. The Enterics for Global Health (EFGH) Shigella Surveillance Study in Bangladesh. Open Forum Infect Dis. 2024;11(Suppl 1):S76–s83.

5. Khan AI, Rashid MM, Islam MT, Afrad MH, Salimuzzaman M, Hegde ST, et al. Epidemiology of Cholera in Bangladesh: Findings From Nationwide Hospital-based Surveillance, 2014-2018. Clinical infectious diseases : an official publication of the Infectious Diseases Society of America. 2020;71(7):1635–42.

6. Seed KD. Battling Phages: How Bacteria Defend against Viral Attack. PLoS Pathog. 2015;11(6):e1004847.

7. Penadés JR, Chen J, Quiles-Puchalt N, Carpena N, Novick RP. Bacteriophage-mediated spread of bacterial virulence genes. Curr Opin Microbiol. 2015;23:171–8.

8. Alam M, Hasan NA, Sadique A, Bhuiyan NA, Ahmed KU, Nusrin S, et al. Seasonal cholera caused by Vibrio cholerae serogroups O1 and O139 in the coastal aquatic environment of Bangladesh. Appl Environ Microbiol. 2006;72(6):4096–104.

9. Koelle K, Pascual M, Yunus M. Pathogen adaptation to seasonal forcing and climate change. Proc Biol Sci. 2005;272(1566):971-7.

10. Faruque SM, Islam MJ, Ahmad QS, Faruque AS, Sack DA, Nair GB, et al. Self-limiting nature of seasonal cholera epidemics: Role of host-mediated amplification of phage. Proceedings of the National Academy of Sciences of the United States of America. 2005;102(17):6119–24.

11. Jensen EC, Schrader HS, Rieland B, Thompson TL, Lee KW, Nickerson KW, et al. Prevalence of broad-host-range lytic bacteriophages of Sphaerotilus natans, Escherichia coli, and Pseudomonas aeruginosa. Appl Environ Microbiol. 1998;64(2):575–80.

12. Hooda Y, Islam S, Kabiraj R, Rahman H, Sarkar H, da Silva KE, et al. Old tools, new applications: Use of environmental bacteriophages for typhoid surveillance and evaluating vaccine impact. PLoS neglected tropical diseases. 2024;18(2):e0011822.

13. Nelson EJ, Grembi JA, Chao DL, Andrews JR, Alexandrova L, Rodriguez PH, et al. Gold Standard Cholera Diagnostics Are Tarnished by Lytic Bacteriophage and Antibiotics. Journal of clinical microbiology. 2020;58(9).

14. Rahman M, Sack DA, Mahmood S, Hossain A. Rapid diagnosis of cholera by coagglutination test using 4-h fecal enrichment cultures. Journal of clinical microbiology. 1987;25(11):2204–6.

15. Qadri F, Das SK, Faruque AS, Fuchs GJ, Albert MJ, Sack RB, et al. Prevalence of toxin types and colonization factors in enterotoxigenic Escherichia coli isolated during a 2-year period from diarrheal patients in Bangladesh. Journal of clinical microbiology. 2000;38(1):27–31.

16. Organization WH. Manual for laboratory investigations of acute enteric infections: programme for control of diarrhoeal diseases: World Health Organization; 1987.

17. Begum YA, Talukder KA, Azmi IJ, Shahnaij M, Sheikh A, Sharmin S, et al. Resistance Pattern and Molecular Characterization of Enterotoxigenic Escherichia coli (ETEC) Strains Isolated in Bangladesh. PloS one. 2016;11(7):e0157415.

18. Waturangi DE. Enumeration of Bacteriophages by Plaque Assay. Methods Mol Biol. 2024;2738:147–53.

19. Daubie V, Chalhoub H, Blasdel B, Dahma H, Merabishvili M, Glonti T, et al. Determination of phage susceptibility as a clinical diagnostic tool: A routine perspective. Frontiers in cellular and infection microbiology. 2022;12:1000721.

20. Qadri F, Svennerholm AM, Faruque AS, Sack RB. Enterotoxigenic Escherichia coli in developing countries: epidemiology, microbiology, clinical features, treatment, and prevention. Clinical microbiology reviews. 2005;18(3):465–83.

21. Tauheed I, Ahmed T, Akter A, Firoj MG, Ahmmed F, Rahman SIA, et al. A snap-shot of a diarrheal epidemic in Dhaka due to enterotoxigenic Escherichia coli and Vibrio cholerae O1 in 2022. Front Public Health. 2023;11:1132927.

22. Alam M, Islam A, Bhuiyan NA, Rahim N, Hossain A, Khan GY, et al. Clonal transmission, dual peak, and off-season cholera in Bangladesh. Infect Ecol Epidemiol. 2011;1.

23. Nelson EJ, Harris JB, Morris JG, Jr., Calderwood SB, Camilli A. Cholera transmission: the host, pathogen and bacteriophage dynamic. Nature reviews Microbiology. 2009;7(10):693–702.

24. Faruque SM, Naser IB, Islam MJ, Faruque AS, Ghosh AN, Nair GB, et al. Seasonal epidemics of cholera inversely correlate with the prevalence of environmental cholera phages. Proceedings of the National Academy of Sciences of the United States of America. 2005;102(5):1702–7.

25. Rogawski McQuade ET, Blake IM, Brennhofer SA, Islam MO, Sony SSS, Rahman T, et al. Real-time sewage surveillance for SARS-CoV-2 in Dhaka, Bangladesh versus clinical COVID-19 surveillance: a longitudinal environmental surveillance study (December, 2019-December, 2021). Lancet Microbe. 2023;4(6):e442–e51.

26. Kim YT, Lee K, Lee H, Son B, Song M, Lee SH, et al. Development of a wastewater based infectious disease surveillance research system in South Korea. Scientific reports. 2024;14(1):24544.

27. Manna T, Chandra Guchhait K, Jana D, Dey S, Karmakar M, Hazra S, et al. Wastewater-based surveillance of Vibrio cholerae: Molecular insights on biofilm regulatory diguanylate cyclases, virulence factors and antibiotic resistance patterns. Microbial pathogenesis. 2024;196:106995.

28. Islam MT, Hegde ST, Khan AI, Bhuiyan MTR, Khan ZH, Ahmmed F, et al. National Hospital-Based Sentinel Surveillance for Cholera in Bangladesh: Epidemiological Results from 2014 to 2021. The American journal of tropical medicine and hygiene. 2023;109(3):575–83.

29. Das R, Nasrin S, Palit P, Sobi RA, Sultana AA, Khan SH, et al. Vibrio cholerae in rural and urban Bangladesh, findings from hospital-based surveillance, 2000-2021. Scientific reports. 2023;13(1):6411.

30. Madi N, Cato ET, Abu Sayeed M, Creasy-Marrazzo A, Cuénod A, Islam K, et al. Phage predation, disease severity, and pathogen genetic diversity in cholera patients. Science. 2024;384(6693):eadj3166.

31. Subramanian S, Parent KN, Doore SM. Ecology, Structure, and Evolution of Shigella Phages. Annu Rev Virol. 2020;7(1):121–41.

32. Ahamed SKT, Rai S, Guin C, Jameela RM, Dam S, Muthuirulandi Sethuvel DP, et al. Characterizations of novel broad-spectrum lytic bacteriophages Sfin-2 and Sfin-6 infecting MDR Shigella spp. with their application on raw chicken to reduce the Shigella load. Frontiers in microbiology. 2023;14:1240570.

33. Goodridge LD. Bacteriophages for managing Shigella in various clinical and non-clinical settings. Bacteriophage. 2013;3(1):e25098.

